# Sex-specific cardiovascular risk factors in the UK Biobank

**DOI:** 10.1101/2023.10.26.23297622

**Authors:** Skyler St. Pierre, Bartosz Kaczmarski, Mathias Peirlinck, Ellen Kuhl

## Abstract

The lack of sex-specific cardiovascular disease criteria contributes to the under-diagnosis of women compared to men. For more than half a century, the Framingham Risk Score has been the gold standard to estimate an individual’s risk of developing cardiovascular disease based on age, sex, cholesterol levels, blood pressure, diabetes, and smoking. Now, machine learning can offer a much more nuanced insight into predicting the risk of cardiovascular disease. The UK Biobank is a large database that includes traditional risk factors as well as tests related to the cardiovascular system: magnetic resonance imaging, pulse wave analysis, electrocardiograms, and carotid ultrasounds. Here we leverage 20,542 datasets from the UK Biobank to build more accurate cardiovascular risk models than the Framingham Risk Score, and quantify the under-diagnosis of women compared to men. Strikingly, for first-degree atrioventricular block and dilated cardiomyopathy, two conditions with non-sex-specific diagnostic criteria, our study shows that women are under-diagnosed 2x and 1.4x more than men. Similarly, our results demonstrate the need for sex-specific criteria in essential primary hypertension and hypertrophic cardiomyopathy. Our feature importance analysis reveals that, out of the top 10 features across three sex and four disease categories, traditional Framingham factors made up between 40-50%, electrocardiogram 30-33%, pulse wave analysis 13-23%, and magnetic resonance imaging and carotid ultrasound 0-10%. Improving the Framingham Risk Score by leveraging big data and machine learning allows us to incorporate a wider range of biomedical data and prediction features, enhance personalization and accuracy, and continuously integrate new data and knowledge, with the ultimate goal to improve accurate prediction, early detection, and early intervention in cardiovascular disease management.

Our analysis pipeline and trained classifiers are freely available at https://github.com/LivingMatterLab/CardiovascularDiseaseClassification

## 1 Motivation

Historically, women have been excluded from the biomedical literature, and clinical and animal trials have been biased towards male-only or male-dominated populations [1]. Including sex as a biological variable is increasingly recognized as essential to decrease health inequities [2, 3]. Sex is typically reported as a binary variable, but sex is inherently complex and relates to hormones, chromosomes, and physical characteristics, all of which follow distributions that overlap between the traditional *male* and *female* categories [4]. In the UK Biobank, sex is reported as a binary variable, and the language used in this study reflects that limitation [5].

### Women are under-diagnosed and under-treated compared to men

Cardiovascular disease is underdiagnosed in women compared to men; the lack of sex-specific diagnostic criteria contributes to this issue [6]. With the currently used, non-sex-specific criteria, the prevalence of dilated and hypertrophic cardiomyopathy is 3:1 and 3:2 men-to-women, indicating that men are diagnosed more frequently than women for these cardiomyopathies [7, 8]. On average, women have a smaller wall thickness than men. For hypertrophic cardiomyopathy, the lack of sex-specific criteria implies that female hearts have to increase in thickness disproportionally more than male hearts to reach the diagnostic threshold of 15mm wall thickness, [9]. Strikingly, women are half as likely to be diagnosed during a routine exam for hypertrophic cardiomyopathy compared to men [7]. Women are also diagnosed at an older age and with more symptoms than men for hypertrophic and dilated cardiomyopathies [7, 8, 10].

The need for sex-specific diagnostic criteria is also visible in heart failure with preserved left ventricle ejection fraction where the cut-off is an ejection fraction of ≥ 50% [11]. However, women have a higher baseline ejection fraction than men on average [12]. In fact, studies have already shown that women benefit from therapies at a higher range of ejection fractions than men [13, 14]. Clearly, there is an urgent need for sex-specific research to understand the different impacts of heart failure on men and women [15–17].

### Risk prediction improve enable early detection

Clinically used risk prediction models for cardiovascular disease typically include components of the Framingham Risk Score: age, sex, total cholesterol, high-density lipoprotein (HDL) cholesterol, systolic blood pressure, blood pressure treated by medicine, diabetes status, and smoking status [18]. Body mass index (BMI) is also common to include in risk models [19]. These risk models are easy to use: They only require a handful of easy-to-measure variables and risk evaluation is a simple score based on discrete thresholds for each of these variables. Clinicians use these risk models to determine if an otherwise asymptomatic person would benefit from a medical intervention [19–22].

### Machine learning models have historically outperformed deep learning for tabular data

Tabular data consists of features that can be put into a spreadsheet, including continuous variables like age and binary variables like smoking status, which are coded with zero for negative and one for positive. The state of the art for supervised learning on tabular data are gradient-boosted tree ensembles [23], which are conventional machine learning methods. The top gradient-boosted models based on benchmark performance for five independent datasets [23] are XGBoost (eXtreme Gradient Boosting) [24], LightGMB [25], and CatBoost [26]. The strengths of tree ensemble methods include robustness against outliers and noisy data [27], and fast exact extraction of feature importance via methods such as SHAP [28]. A weakness of decision trees is that they are unstable and tend to overfit the training data [27].

While gradient boosting frameworks have shown to be the best performing approaches for tabular data in the past decade [29], deep learning architectures are becoming increasingly prevalent [30], sometimes outperforming the state-of-the-art approaches [23, 31]. In particular, deep learning frameworks such as TabTransformer [32], DeepFM [33], TabNet [34], and SAINT (Self-Attention and Intersample Attention Transformer) [31] showed significant promise for effective tabular data modeling, with the SAINT model outperforming gradient-boosted frameworks on some learning tasks by using the power of representation learning [23, 31]. Nonetheless, a well-known downside of deep learning models, as compared to tree-based frameworks, is that they are generally much slower to train [35].

### Machine learning can discover the most predictive features for risk models

Machine learning models can effectively utilize a large number of input features, which allows them to discover new, more accurate risk models to better identify people at risk [19, 36, 37]. XGBoost has been applied extensively for cardiovascular disease diagnosis [19, 38–41]. Prior statistical or deep learning models of cardiovascular disease focused on lifestyle factors [19, 22, 41, 42], medical history [19, 41], sociodemographics [19, 41], dietary and nutritional information [19, 41], genetics [41, 42], and/or one of four clinical tests: pulse wave analysis [43, 44], electro-cardiogram [41, 45, 46], carotid ultrasound [42, 47], or magnetic resonance imaging [48–50], but not all four.

### Objectives of this study

First, we investigate whether women are under-diagnosed for cardiovascular diseases in the UK Biobank cohort. Then, we compare the performance of three models, a multilayer perceptron deep learning baseline, XGBoost, and the novel deep learning framework, SAINT, on their ability to predict whether a person can be diagnosed with a cardiovascular disease. Lastly, we identify the top sex- and disease-specific risk factors from four cardiovascular-related tests, pulse wave analysis, electrocardiograms, magnetic resonance imaging, and carotid ultrasound, against the traditional Framingham Risk Score.

## 2 Methods

### 2.1 Dataset and features

The UK Biobank is comprised of data from half a million individuals from the UK over the age of 40 [5]. From these, we selected individuals who underwent ECG testing, magnetic resonance imaging, carotid ultrasounds, and pulse wave analysis resulting in a population of 20,542 individuals. We also pulled features associated with the Framingham Risk Score, sex, age, total cholesterol, HDL cholesterol, smoking status, diabetes status, end systolic blood pressure, the body mass index of all participants, and their medical diagnoses. We did not include treatment for blood pressure as a feature in our models as this directly reflects one of the diagnostic outcomes, hypertension, that we are trying to predict.

We created two feature groups: i) 8 features including Framingham Risk Score features and body mass index only, and ii) all 57 features. We labeled each person to be in the positive class if they were diagnosed with a given cardiovascular disease [43]. We split the datasets based on four disease categories: any disease, hypertension (ICD10 codes I10-I15), ischemic (I20-I25), and conduction disorders (I44-I49) [51], such that the detection of each disease can be posed as a binary classification. To train the sex-specific classifiers, we further designated three input groups, i.e., both sexes, female only, and male only.

Using the three input groups (both sexes, female only, male only) together with the four binary label sets (any disease, hypertensive disease, ischemic heart disease, conduction disorders), we constructed 12 dataset variants to train the binary classifiers, see Figure 1. We generated each of the datasets by direct slicing of the randomly pre-shuffled dataframe. Since the datasets are relatively small, we applied a 70-15-15 split to create the training, validation, and test sets for each of the variants. The positive class in all 12 dataset variants is significantly underrepresented relative to the negative class, so we applied oversampling to approximately equalize the number of negative and positive samples in the training sets of the corresponding 12 datasets.

**Fig. 1.**
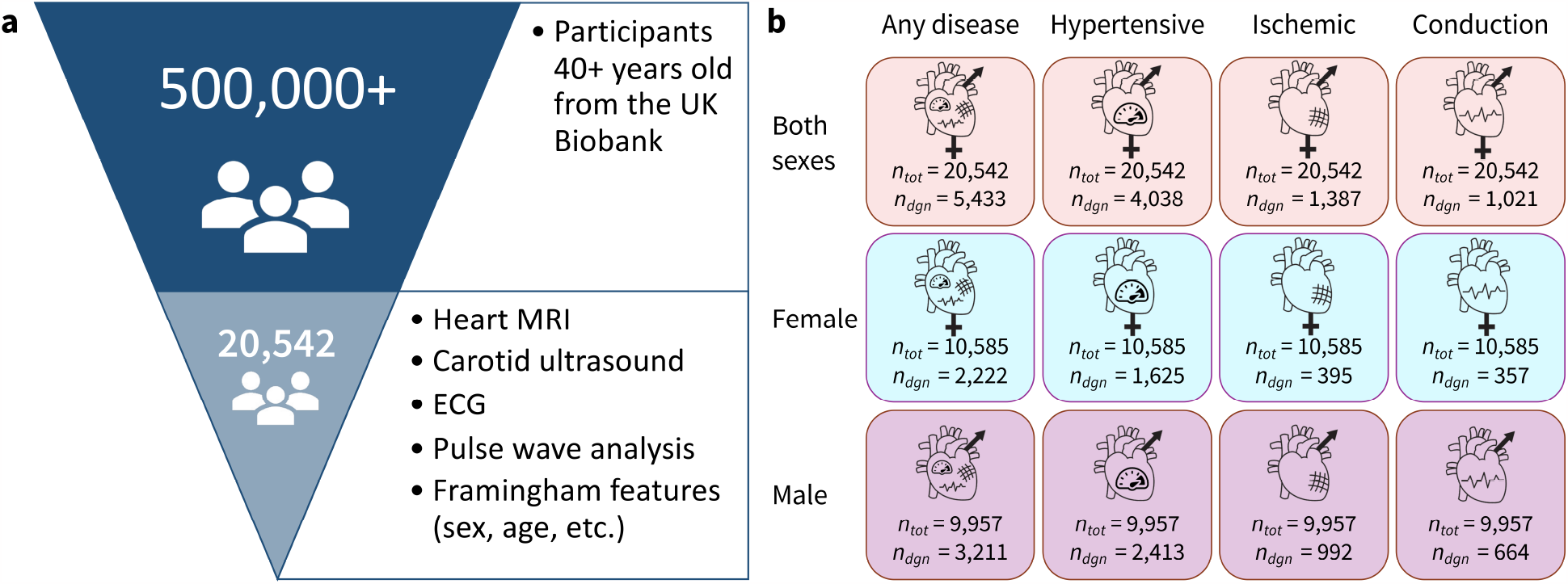
Dataset overview. (a) Out of 500,000+ participants in the UK Biobank study we selected a group of 20,542 participants who underwent magnetic resonance imaging, carotid ultrasound, ECG, and pulse wave analysis. We also selected participants with available data for all of the Framingham Risk Score features. (b) The data are separated in 12 variants with three sex groups and four cardiovascular disease categories where *n*_*tot*_ is the total number of people in the dataset and *n*_*diag*_ is the number of people in that dataset who have been diagnosed with the corresponding condition.

### 2.2 Models

Using the cardiovascular and Framingham Risk Score features, we implemented three distinct model types: i) a multilayer preceptron (MLP), ii) an XGBoost ensemble model, which is the state of the art approach for tabular data learning [24], and iii) the SAINT model [31]. We used the MLP as a baseline for deep-learning performance and the XGBoost as a baseline for state of the art performance. For each model type, we trained and evaluated 12 individual classifiers according to our 12 dataset variants. For evaluation purposes, we consider both untuned and tuned XGBoost ensemble models, and introduce an additional set of tuned XGBoost ensembles trained only on the Framingham Risk Score features. The 12 cardiovascular disease datasets, three model types, and two additional XGBoost variants result in a total of 60 individually trainable classifiers.

#### MLP: A deep learning model for baseline comparison

We implemented and evaluated a baseline multilayer perceptron (MLP) network under TensorFlow [52] for each of the 12 dataset variants. All 12 MLP classifiers were trained using the binary cross-entropy function with *L*_2_-regularization in the cost. To accelerate and stabilize training, the features are first passed through a standarization layer, and each hidden layer is followed by a batch normalization layer. Each layer uses a ReLU non-linearity and the output uses a sigmoid activation function. The tunable hyperparameters of the MLP are the number of hidden layers, the number of units in each hidden layer, the *L*_2_-regularization parameter, and the parameters of the training procedure.

#### XGBoost: A state of the art analysis of tabular data

We used XGBoost [24] as a benchmarking baseline for the novel SAINT model. XGBoost trains an ensemble of decision tree models using an efficient second-order gradient boosting framework [24]. We trained an individual XGBoost ensemble for each of the 12 dataset variants using the binary cross entropy loss and training-test splits consistent with the ones used for the MLP models.

#### SAINT: A novel approach for tabular data learning

The Self-Attention and Intersample Attention Transformer (SAINT) is a novel approach for tabular data modeling that employs self-attention, intersample attention, an enhanced embedding framework, and a contrastive pre-training phase [31]. Transformers are a recent development in neural network research employed in cutting-edge generative machine learning applications such as ChatGPT-4 [53]. They have been shown to significantly outperform previous state-of-the-art machine learning architectures for language modeling and machine translation tasks [54]. Rather than using transformers for language processing purposes, SAINT adapts this architecture and the concept of self-attention to perform efficient learning on tabular data, such as the clinical UK Biobank data analyzed in our study. While SAINT has been shown to outperform the state of the art methods on some datasets [23], it has not yet been used for cardiovascular data learning. Here we applied SAINT to investigate its performance in cardiovascular disease classification tasks, in addition to the more established MLP and XGBoost methods.

### 2.3 Model evaluation

#### ROC: Receiver operating characteristic curve

The receiver operating characteristic (ROC) curve is used to evaluate the performance of a diagnostic test where the predictors of the outcome are not binary, so there are many possible cut-points to classify a person with a positive or negative diagnosis [55]. The ROC curve is a plot of sensitivity (true positive rate) vs. 1-specificity (false positive rate). Sensitivity is the probability that an individual who is truly positive gets a positive test result, while specificity is the probability that an individual who is truly negative gets a negative test result [56]. The diagonal line indicates that whether or not a person is diagnosed is totally random.

#### AUC: Area under the curve

The area under the curve (AUC) is a summary metric for the ROC curve that reports the overall accuracy of the test [55]. The AUC ranges from 0, completely inaccurate, to 1, completely accurate, with an AUC of 0.5 meaning that the test result is random. We use the ROC curve and AUC metric to compare how accurately our different models can predict cardiovascular disease, as the ROC curve does not depend on the scale of the test results and provides a helpful visual comparison [55].

#### Feature importance rankings

SHapley Additive exPlanations (SHAP) is a unified framework designed to interpret model predictions by giving a value for the importance of each feature to a specific prediction [28]. A positive SHAP value indicates that a feature has a positive impact on the prediction of the positive class, in our case, a diagnosis of a cardiovascular disease, while a negative SHAP values indicates the opposite. The magnitude indicates the strength of the effect. We can easily integrate the SHAP pipeline with XGBoost using the TreeExplainer class.

## 3 Results

### Women are under-diagnosed relative to men

We first investigate whether women are under-diagnosed relative to men for cardiovascular diseases in the UK Biobank cohort. We chose diseases where the diagnosis is a simple, non-sex-specific cut-off.

Figure 2 shows the cut-off criteria in red. Plots (a-d) show indivdiuals who have not been diagnosed with the disease in orange, and those who have in purple. Plots (e,f) are divided into four categories. The truncated violin plots show the distribution of each sex for each category with the box-plots showing the mean in white and the 25th-75th percentiles. Each dot represents a single person in the Biobank dataset. There may be co-morbidities or alternate medical diagnoses that result in similar presentations so the magnitude of under-diagnosis in the following examples should be understood as a first approximation.

**Fig. 2.**
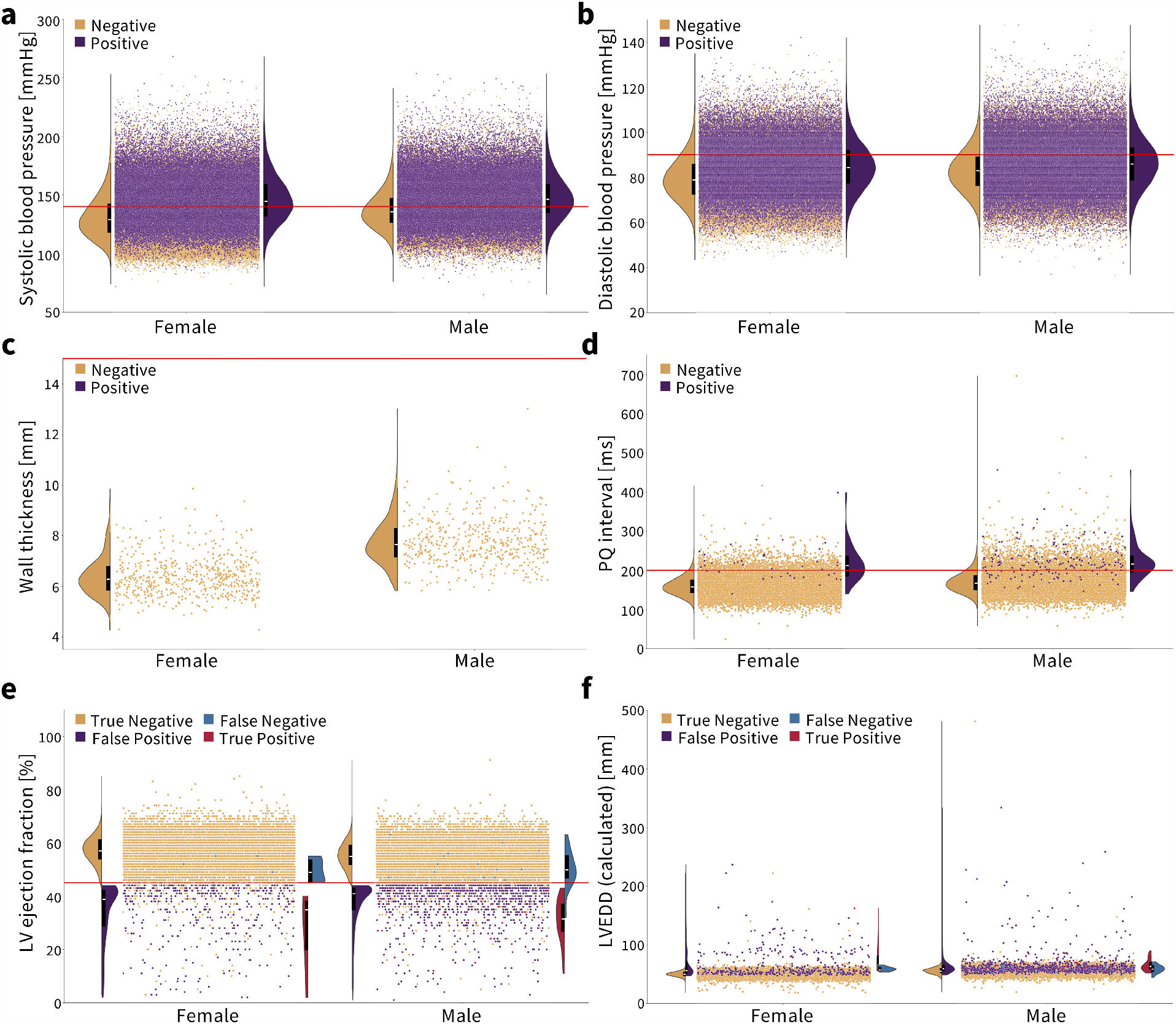
Diagnosing cardiovascular disease via simple, non-sex-specific cut-offs. The red line indicates the diagnostic cut-off. The truncated violin plots show the distribution of men and women for each color-coded population, with the box-plot inside showing the mean in white and the 25th and 75th percentiles. (a, b) Essential primary hypertension is diagnosed with a systolic blood pressure greater than or equal to 140 mmHg and/or diastolic blood pressure greater than or equal to 90 mmHg [57]. Women who are not diagnosed with hypertension have on average a lower systolic and diastolic blood pressure compared to men. (c) Hypertrophic cardiomyopathy is diagnosed with a wall thickness greater than 15 mm [58]. None of the individuals in this cohort met the condition. Healthy women have a notably lower wall thickness on average than men. (d) First degree AV block is diagnosed with a PQ interval greater than 200 ms [59]. Healthy women have a lower PQ interval on average than men. (e, f) Dilated cardiomyopathy is diagnosed by a left ventricle ejection fraction less than 45% and a left ventricle end-diastolic diameter greater than 112% of the diameter predicted based on body surface area and age [60, 61]. In orange, women have a slightly higher ejection fraction and lower left ventricle end diastolic diameter on average than men.

*Essential primary hypertension* is diagnosed by a systolic blood pressure ≥ 140 mmHg and/or diastolic blood pressure ≥ 90 mmHg [57]. The corresponding ICD10 code is I10. Figure 2(a, b) show that women have on average a lower systolic and diastolic blood pressure than men. In the Biobank cohort, 35.2% of men are diagnosed while 52.3% of men meet the cut-off criteria. For women, 26.6% are diagnosed while 40.2% meet the cut-off criteria. This means that women and men are under-diagnosed for essential primary hypertension at the same rate, 1.5x, when non-sex-specific criteria are used.

*Hypertrophic cardiomyopathy* is diagnosed with a wall thickness *>* 15 mm [58]; the ICD10 codes are I42.1 and I42.2. Figure 2(c) shows that none of the approximately 900 people with cardiac magnetic resonance images met the criteria for hypertrophic cardiomyopathy or were diagnosed. Women have a distinctly smaller wall thickness on average than men.

*First degree AV block* is diagnosed with a PQ interval *>* 200 ms [59]; the ICD10 code is I44.0. As shown in Figure 2(d), women have a smaller PQ interval than men on average. First degree AV block is generally asymptomatic but no longer consider entirely benign, with nearly double the risk of developing atrial fibrillation and triple the risk of needing a pacemaker [59]. As such, the current recommendation is to monitor patients regularly to see if the conduction delay continues to widen or if they are developing atrial fibrillation [62]. In the UK Biobank, 0.81% of men are diagnosed and 12.6% of men meet the cut-off. For women, 0.18% are diagnosed while 5.4% meet the cut-off. So, women are under-diagnosed 30x while men are under-diagnosed 15.6x, meaning that women are nearly 2x more under-diagnosed relative to men for first degree AV block given non-sex-specific criteria.

*Dilated cardiomyopathy* is diagnosed by a left ventricle ejection fraction *<* 45% and a left ventricle end-diastolic diameter *>* 112% of the predicted diameter based on age and sex [60]. Left ventricle fractional shortening less than 25% can be used in place of the ejection fraction criteria, but these data were not available in the Biobank. Because left ventricle end-diastolic volume is reported, we used the Teichholz formula [61], LVEDV = 7(LVEDD_cal_)^3^*/*(2.4 + LVEDD_cal_), to calculate the end-diastolic diameter from the volume, and the formula, LVEDD_pre_ = 45.3(BSA)^0.3^ − 0.03(age) − 7.2, to predicted the end-diastolic diameter from BSA and age. If LVEDD_cal_*/*LVEDD_pre_ *>* 1.12, the individual would meet the criteria and be assigned either a red or purple dot in Figure 2(e,f), depending on whether they had also been diagnosed with dilated cardiomyopathy or not, respectively. If they did not meet this criterion, they were assigned an orange or blue dot, where blue indicates they had been diagnosed and orange they had not been. In the Biobank cohort, 55 people were diagnosed with dilated cardiomyopathy, but only 35 met the cut-off using these calculations, with nearly all of these discrepancies for not meeting the ejection fraction criteria as shown in blue. Women have on average a slightly lower end-diastolic diameter and higher ejection fraction than men. Out of the men in the cohort, 0.23% were diagnosed while 5.71% met the cut-off criterion. For women, 0.06% were diagnosed while 2.02% met the cut-off criterion. As such, women are under-diagnosed 33.7x, men are under-diagnosed 24.8x, and women are 1.4x more under-diagnosed than men when non-sex-specific criteria are used.

### The SAINT model performs the best in predicting risk for cardiovascular disease

Figure 3 shows the receiver operator curve (ROC) and area under the curve (AUC) values for the five models types, MLP, untuned XGBoost, tuned XGBoost, SAINT, and XGBoost with only Framingham Risk Score features across the twelve sex and disease categories. A large AUC score is designed to minimize false negatives (predicted healthy but actually diseased) and maximize true positives (predicted diseased and actually diseased). Table 3 summarizes the performance metrics for all classifiers, where we report test set accuracy, precision, and recall in addition to the AUC score. In terms of the AUC metric, the SAINT model performed best on all datasets except the female only conduction disorder datasets, where only the corresponding tuned XGBoost model performed better. For accuracy and precision, the XGBoost (tuned) models were the best performing in 11/12 cases and 8/12 cases. SAINT had the best performing recall in 9/12 cases.

**Fig. 3.**
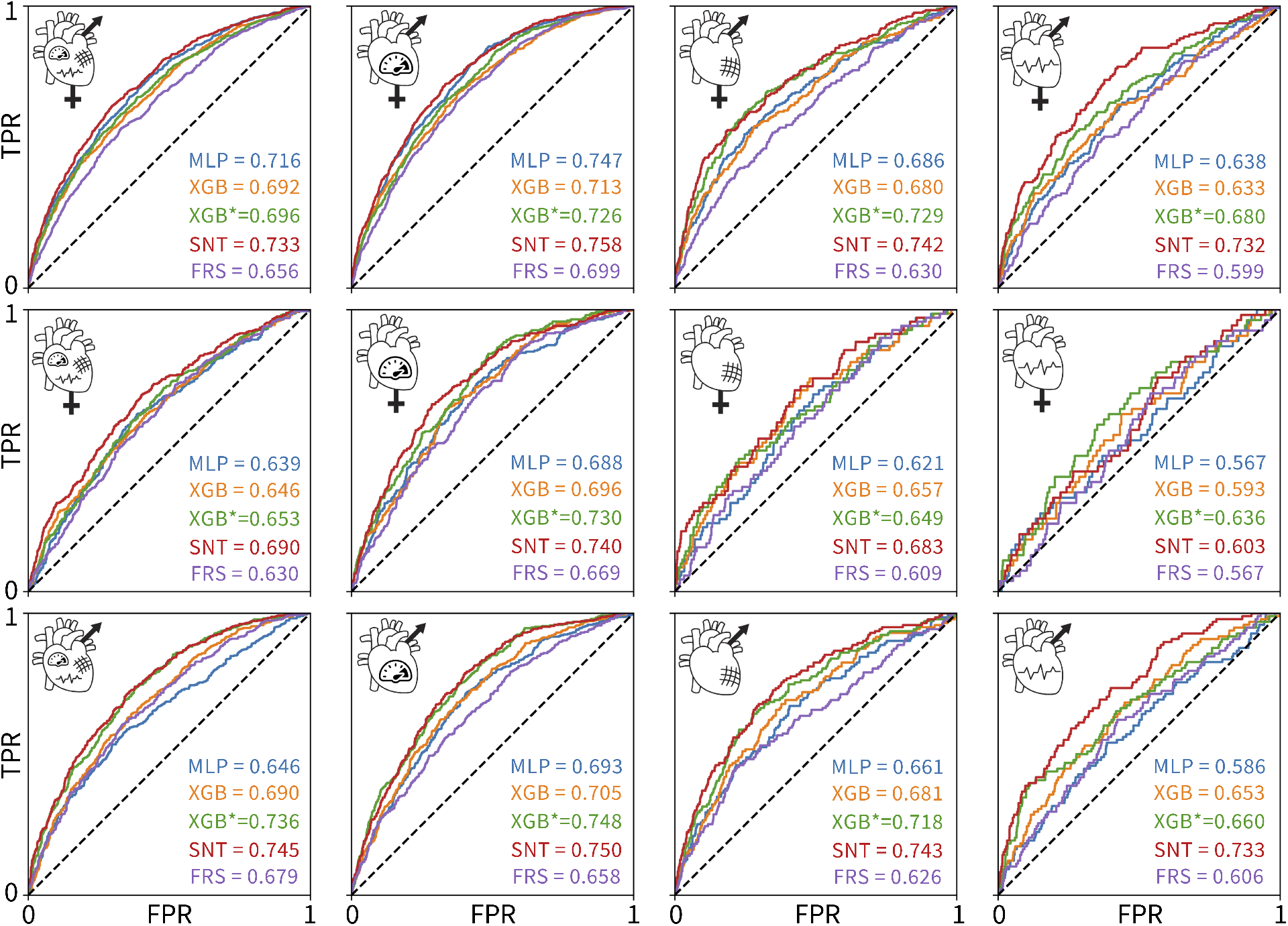
ROC curves and AUC scores for the 60 classifiers evaluated on 12 test sets. The rows correspond to (1) both sexes, (2) female only, and (3) male only datasets. The columns correspond to the (1) any, (2) hypertensive, (3) ischemic, and (4) conduction diseases. The colors of the curves indicate the different model types: MLP deep-learning baseline (blue), untuned XGBoost (orange), tuned XGBoost baseline for state of the art (green), SAINT (red), and XGBoost trained and tuned on Framingham Risk Score features only (purple). True positive rate is plotted versus false positive rate.

While all 60 models were measurably better than a random classifier, none of the models demonstrated a high AUC score. Since we did not observe training set overfitting, this might be indicative of a high Bayes error rate and low feature-output correlations in the datasets. The ischemic disease and conduction disorder models performed rather poorly, most likely caused by the small training set sizes and the increasingly significant class imbalance in their test sets.

### Additional features improves cardiovascular disease prediction

Figure 3 suggests that including features from ECG, magnetic resonance imaging, pulse wave analysis, and carotid ultrasound, along with Framingham Risk Score features significantly increased the AUC score of the corresponding 48 models, as compared to the AUC scores for the Framingham-only XGBoost models. The XGBoost classifiers trained and tuned on the Framingham-only features were always the lowest or second-lowest performing models for a given dataset.

### Predicting the risk of cardiovascular disease for women is less accurate than for men

Figure 4 shows the performance of the twelve individually trained XGBoost classifiers on individual sexes. First, the classifiers trained on both sexes perform the best for all female only datasets, top row. The best AUC values for the female only data are lower than the best AUC values for the male only data for all disease categories except conduction disorders. Second, the male-only classifiers perform the best for the male-only datasets for any-disease and hypertension categories, while the both-sexes classifiers perform the best for the ischemic and conduction diseases. Third, the performance of all classifiers is fairly similar for most sex- and disease-specific categories except for three cases: i) the female-only classifier is significantly worse at predicting male cases of ischemic diseases, ii) the male-only classifier is worse at predicting female cases of ischemic diseases, and iii) at predicting conduction diseases as compared to the female-only and both-sexes classifiers.

**Fig. 4.**
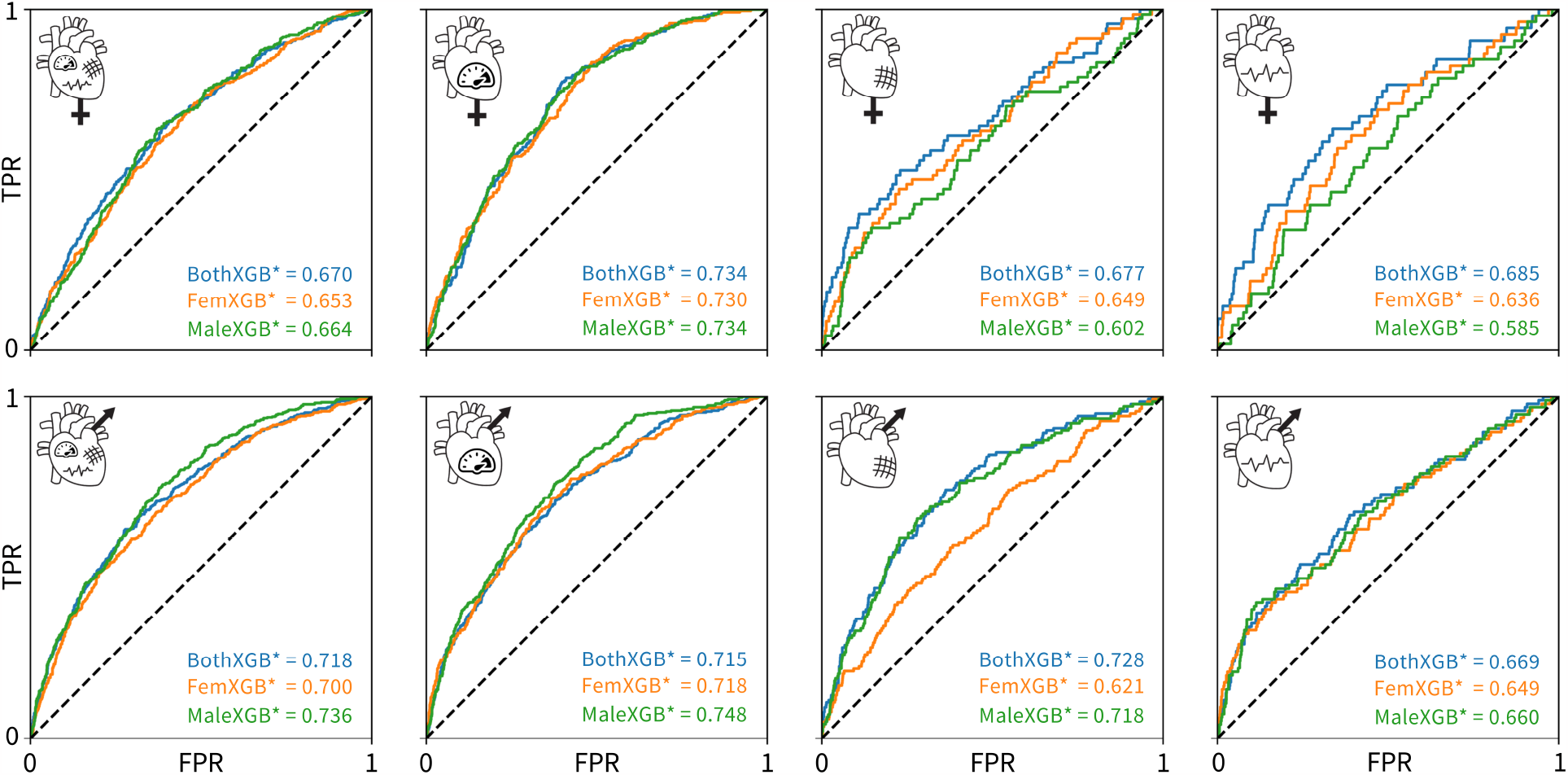
Cross-evaluation results using the tuned XGBoost classifiers. The classifiers trained on both sexes are colored blue, the classifiers trained on only female data are colored orange, and the classifiers trained on only male data are colored green. The rows show the ROC and AUC for a given trained classifier in predicting a given disease for only female data, top, or only male data, bottom. The columns correspond to any cardiovascular disease, hypertensive diseases, ischemic diseases, and conduction diseases, from left to right. True positive rate is plotted versus false positive rate.

### A subset of Framingham Risk Score and ECG features are most predictive for cardiovascular disease

Figure 5 shows the ten most predictive features for any type of cardiovascular disease for both sexes combined, women only, and men only. A more positive SHAP value indicates a larger contribution to the positive class, diagnosed with a cardiovascular disease, while a negative SHAP value indicates the opposite. Each dot represents an individual person in the dataset, while red color means that person had a high value of that feature, e.g., older while blue means a lower value, e.g., younger. For the binary categories of sex, smoking status, and diabetes status, red means male, a person who smokes, and a person with diabetes. Traditional risk factors refer to the Framingham Risk Score features plus body mass index as shown in Table 1.

**Table 1.** Features for inclusion in risk prediction analysis. The traditional Framingham Risk Score and body mass index are commonly used factors in clinical risk prediction for cardiovascular disease. Features from magnetic resonance imaging, carotid ultrasound, electrocardiogram recordings, and pulse wave analysis are also extracted from the UK Biobank.

| Method | Features |
| --- | --- |
| Framingham Risk Score + body mass index (8) | Sex, age, high-density lipoprotein cholesterol, total cholesterol, end systolic blood pressure, smoking status, diabetes status, body mass index |
| Magnetic resonance imaging (7) | Average heart rate, cardiac index, cardiac output, left ventricle ejection fraction, left ventricle end-diastolic volume, left ventricle end-systolic volume, left ventricle stroke volume |
| Carotid ultrasound (12) | Max/mean/min carotid intima-media thickness 120/150/210/240° |
| Electrocardiogram (12) | Ventricular rate, P duration, PP interval, PQ interval, QRS number, QRS duration, QT interval, QTC interval, RR interval, P axis, R axis, T axis |
| Pulse wave analysis (19) | Position of pulse wave notch, position of pulse wave peak, position of shoulder on pulse waveform, pulse rate, pulse wave arterial stiffness index, pulse wave peak to peak time, pulse wave reflection index, augmentation index, central augmentation pressure, central pulse pressure, central systolic blood pressure, diastolic blood pressure, end-systolic pressure index, mean arterial pressure index, number of beats in waveform average, peripheral pulse pressure, stroke volume, systolic brachial blood pressure |

**Table 2.** Disease classification criteria. Clinical diagnostic ICD10 codes for different subsets of cardiovascular disease [43].

| Disease | ICD10 Codes |
| --- | --- |
| Any cardiovascular disease | I00-I78.9, G95.1, H334.1-2, O10.0-9, S06.60-61, Z95.1, Z95.5 |
| Hypertensive diseases | I10-I15.9 |
| Ischemic diseases | I20-I25.9 |
| Conduction disorders | I44-I49.9 |

**Table 3.** Comparison of 60 classifiers. For each test set, we report the accuracy (Acc), precision (Prec), recall (Rec), and area under the curve (AUC) scores for each of the five evaluated model types: MLP, untuned XGBoost, tuned XGBoost, SAINT, and XGBoost trained and tuned with Framingham Risk Score features only. Bolded and underlined quantities correspond to the best and second-best values for a given metric and dataset.

|  |  | MLP | XGBoost<br>(untuned) | XGBoost<br>(tuned) | SAINT | XGBoost<br>(Fram. only) |
| --- | --- | --- | --- | --- | --- | --- |
| Both sexes,<br>Any disease | Acc: | 0.652 | 0.704 | **0.732** | 0.682 | <u>0.707</u> |
|  | Prec: | 0.426 | <u>0.473</u> | **0.551** | 0.455 | <u>0.469</u> |
|  | Rec: | **0.667** | <u>0.450</u> | 0.268 | <u>0.651</u> | 0.291 |
|  | AUC: | <u>0.716</u> | 0.692 | 0.696 | **0.733** | 0.656 |
| Both sexes,<br>Hypertension | Acc: | 0.663 | 0.747 | **0.786** | 0.633 | <u>0.767</u> |
|  | Prec: | 0.354 | 0.410 | **0.498** | 0.343 | <u>0.417</u> |
|  | Rec: | <u>0.695</u> | 0.420 | 0.212 | **0.780** | <u>0.226</u> |
|  | AUC: | <u>0.747</u> | 0.713 | 0.726 | **0.758** | 0.699 |
| Both sexes,<br>Ischemic | Acc: | 0.765 | 0.906 | **0.931** | 0.677 | <u>0.919</u> |
|  | Prec: | 0.147 | <u>0.243</u> | **0.542** | 0.137 | <u>0.207</u> |
|  | Rec: | <u>0.491</u> | <u>0.162</u> | 0.060 | **0.681** | 0.056 |
|  | AUC: | <u>0.686</u> | 0.680 | <u>0.729</u> | **0.742** | 0.630 |
| Both sexes,<br>Conduction | Acc: | 0.765 | 0.934 | **0.952** | 0.739 | <u>0.946</u> |
|  | Prec: | 0.085 | <u>0.184</u> | **0.500** | 0.101 | <u>0.050</u> |
|  | Rec: | <u>0.396</u> | <u>0.107</u> | 0.027 | **0.557** | 0.007 |
|  | AUC: | <u>0.638</u> | 0.633 | <u>0.680</u> | **0.732** | 0.599 |
| Female,<br>Any disease | Acc: | 0.634 | <u>0.741</u> | **0.761** | 0.692 | 0.734 |
|  | Prec: | 0.336 | <u>0.417</u> | **0.462** | 0.378 | 0.341 |
|  | Rec: | **0.591** | <u>0.287</u> | 0.182 | <u>0.504</u> | 0.157 |
|  | AUC: | 0.639 | 0.646 | <u>0.653</u> | **0.690** | 0.630 |
| Female,<br>Hypertension | Acc: | 0.716 | 0.797 | **0.837** | 0.770 | <u>0.799</u> |
|  | Prec: | 0.283 | 0.335 | **0.527** | <u>0.354</u> | <u>0.298</u> |
|  | Rec: | <u>0.469</u> | 0.237 | 0.111 | **0.477** | 0.160 |
|  | AUC: | <u>0.688</u> | 0.696 | <u>0.730</u> | **0.740** | 0.669 |
| Female,<br>Ischemic | Acc: | 0.857 | **0.955** | <u>0.951</u> | 0.700 | 0.948 |
|  | Prec: | 0.089 | **0.333** | <u>0.167</u> | 0.075 | 0.000 |
|  | Rec: | <u>0.243</u> | 0.029 | <u>0.029</u> | **0.514** | 0.000 |
|  | AUC: | <u>0.621</u> | <u>0.657</u> | 0.649 | **0.683** | 0.609 |
| Female,<br>Conduction | Acc: | 0.880 | <u>0.965</u> | **0.965** | 0.945 | 0.964 |
|  | Prec: | 0.070 | **0.250** | 0.000 | <u>0.075</u> | 0.000 |
|  | Rec: | **0.204** | 0.019 | 0.000 | <u>0.056</u> | 0.000 |
|  | AUC: | 0.567 | 0.593 | **0.636** | <u>0.603</u> | 0.567 |
| Male,<br>Any disease | Acc: | 0.603 | 0.675 | **0.709** | 0.669 | <u>0.687</u> |
|  | Prec: | 0.434 | 0.518 | **0.641** | 0.505 | <u>0.552</u> |
|  | Rec: | <u>0.609</u> | 0.426 | 0.296 | **0.668** | <u>0.348</u> |
|  | AUC: | <u>0.646</u> | 0.690 | <u>0.736</u> | **0.745** | 0.679 |
| Male,<br>Hypertension | Acc: | 0.662 | 0.718 | **0.755** | 0.690 | <u>0.722</u> |
|  | Prec: | 0.406 | 0.459 | **0.603** | 0.444 | <u>0.461</u> |
|  | Rec: | <u>0.589</u> | 0.356 | 0.222 | **0.677** | <u>0.285</u> |
|  | AUC: | <u>0.693</u> | 0.705 | <u>0.748</u> | **0.750** | 0.658 |
| Male,<br>Ischemic | Acc: | 0.723 | 0.878 | **0.893** | 0.751 | <u>0.890</u> |
|  | Prec: | 0.167 | <u>0.262</u> | 0.257 | 0.209 | **0.267** |
|  | Rec: | <u>0.476</u> | <u>0.154</u> | 0.063 | **0.573** | 0.084 |
|  | AUC: | <u>0.661</u> | 0.681 | <u>0.718</u> | **0.743** | 0.626 |
| Male,<br>Conduction | Acc: | 0.807 | <u>0.925</u> | **0.932** | 0.612 | 0.922 |
|  | Prec: | 0.100 | **0.281** | <u>0.250</u> | 0.114 | 0.087 |
|  | Rec: | <u>0.245</u> | 0.092 | <u>0.020</u> | **0.724** | 0.020 |
|  | AUC: | <u>0.586</u> | 0.653 | <u>0.660</u> | **0.733** | 0.606 |

**Fig. 5.**
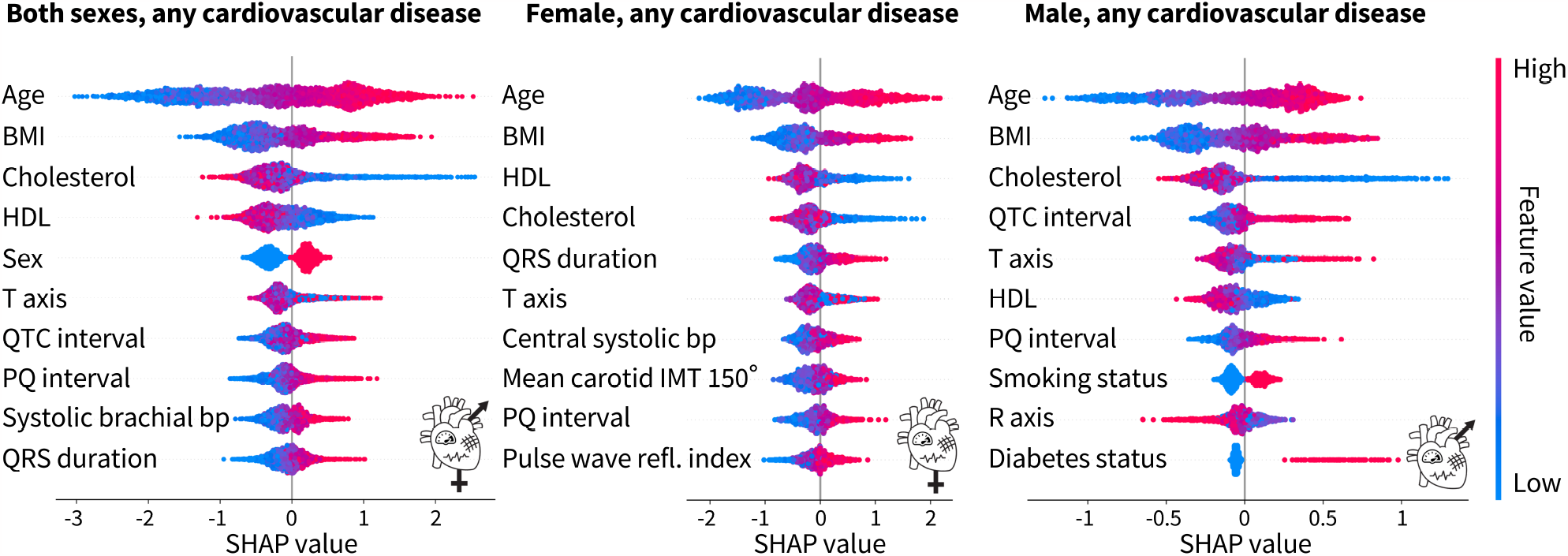
Top 10 features from the tuned XGBoost classifiers trained on both sexes, female only, and male only for any cardiovascular disease. For both sexes, the top four features for the prediction of cardiovascular disease are traditional risk factors, while ECG features and a blood pressure feature from pulse wave analysis make up the rest of the top ten. For the female only dataset, in addition to the top four traditional risk factors, there is a mix of ECG, pulse wave, and carotid ultrasound features. For the male only dataset, six of the features are traditional risk factors while the rest are ECG features. Each dot corresponds to a person in the SHAP analysis dataset. A positive SHAP value indicates contribution to a diagnosis of cardiovascular disease. Bright red corresponds to a high feature value, e.g. old age, while bright blue corresponds to low feature value, e.g. young age. The binary categories of sex, smoking status, and diabetes status are red for male, smoker, and diabetic, while blue is the opposite.

For both sexes combined, six of the top ten features, age, body mass index, cholesterol, HDL cholesterol, sex, and blood pressure are factors traditionally associated with increased cardiovascular risk. The other four features are ECG features. For the female-only dataset, five of the top ten features are traditional risk factors but the rest of the features include a mix of ECG, pulse wave analysis, and carotid ultrasound features. For the male-only dataset, six of the traditional risk factors make the top ten features. The other four features are ECG features. Interestingly, the male-only dataset is the only place where smoking status and diabetes status make the top ten features. Age, body mass index, and cholesterol, either HDL or total, are consistently the top three features regardless of sex. The ECG features of PQ interval and T axis appear in all three categories as well.

Table 4 shows the top ten features based on the SHAP value for the tuned XGBoost model prediction of cardiovascular disease. Across all groups, age is the most important feature in predicting risk. When trained on both sexes, body mass index and HDL cholesterol also appear for all disease groups. Sex is the most important for ischemic heart disease, but interestingly is not in the top ten for conduction disorders. Out of the traditional risk factors in the Framingham Risk Score, diabetes status appears only once for the prediction of hypertension, and smoking status not at all. A measure of blood pressure also only appears for any disease and hypertension. The ECG, pulse wave analysis, and magnetic resonance imaging features of PQ interval, T axis, pulse rate, R axis, QRS duration, and LV ejection fraction also appear in two disease categories each in the top ten.

**Table 4.** Top 10 features to predict of cardiovascular disease for each sex and disease group. Rankings are reported using SHAP on the XGBoost classifiers trained and tuned on each of the 12 datasets with all features from Table 1. BMI body mass index, BP blood pressure, HDL high density lipoprotein, IM intima media, LV left ventricle, PW pulse wave.

|  | Any disease | Hypertension | Ischemic | Conduction |
| --- | --- | --- | --- | --- |
| Both sexes | Age<br>BMI<br>Cholesterol<br>HDL<br>Sex<br>T axis<br>QTC interval<br>PQ interval<br>Systolic brachial BP<br>QRS duration | Age<br>BMI<br>Cholesterol<br>HDL<br>Central systolic BP<br>Sex<br>R axis<br>Mean arterial pressure<br>Diabetes status<br>QTC interval | Age<br>HDL<br>Sex<br>Cholesterol<br>Pulse rate<br>T axis<br>BMI<br>LV ejection fraction<br>R axis<br>LV end systolic vol. | Age<br>PQ interval<br>QRS duration<br>T axis<br>QTC interval<br>LV end systolic vol.<br>P duration<br>HDL<br>BMI<br>Pulse rate |
| Female only | Age<br>BMI<br>HDL<br>Cholesterol<br>QRS duration<br>T axis<br>Central systolic BP<br>Mean carotid IM thickness 150°<br>PQ interval<br>PW reflection index | Age<br>BMI<br>Central systolic BP<br>HDL<br>Mean arterial pressure<br>R axis<br>Cholesterol<br>QRS duration<br>Systolic brachial BP<br>End systolic BP | Age<br>HDL<br>Max carotid IM thickness 240°<br>Cholesterol<br>T axis<br>BMI<br>Pulse rate<br>PW arterial stiff. index<br>Mean carotid IM thickness 120°<br>Mean carotid IM thickness 210° | Age<br>HDL<br>QTC interval<br>P duration<br>T axis<br>QRS duration<br>Cent. augment. press.<br>PQ interval<br>Augmentation index<br>P axis |
| Male only | Age<br>BMI<br>Cholesterol<br>QTC interval<br>T axis<br>HDL<br>PQ interval<br>Smoking status<br>R axis<br>Diabetes status | Age<br>BMI<br>Cholesterol<br>Central systolic BP<br>QTC interval<br>Systolic brachial BP<br>Diabetes status<br>R axis<br>PQ interval<br>Cent. augment. press. | Age<br>Cholesterol<br>HDL<br>PW stroke volume<br>Pulse rate<br>T axis<br>BMI<br>LV stroke volume<br>Max carotid IM thickness 210°<br>Diastolic BP | Age<br>T axis<br>QTC interval<br>PQ interval<br>QRS duration<br>PW stroke volume<br>BMI<br>Pulse rate<br>LV end systolic vol.<br>P duration |

#### Female-only dataset

HDL cholesterol is the only other feature to appear in all categories while body mass index, total cholesterol, and T axis appear three times. Smoking status and diabetes status do not appear at all. Hypertension is heavily predicted by four different measures of blood pressure, while ischemic heart disease is predicted by several measures of intima-media-thickness from carotid ultrasound. Lastly, the ECG feature PQ interval appears in two categories.

#### Male-only dataset

Body mass index is the only other feature to appear in all categories while T axis, total cholesterol and QTC interval appear in three of the disease categories. Six of the features for any disease are traditional risk factors. The other four are ECG features. For hypertension, blood pressure measures and diabetes status along with traditional risk factors like age, body mass index, and cholesterol contribute to the risk prediction. Interestingly, three ECG features also make up the top ten features. For ischemic diseases, stroke volume and a carotid ultrasound feature add to the traditional risk factors of age, cholesterol, HDL cholesterol, body mass index, and blood pressure. Pulse rate and T axis round out the top ten. For conduction disorders, five of the features are ECG features. Age and body mass index are the only traditional risk factors. Magnetic resonance imaging and pulse wave analysis features of pulse rate, stroke volume, and LV end systolic volume are the rest of the top ten.

#### Both sexes combined

Traditional risk factors make up 47.5% of the top ten features, while magnetic resonance imaging features are 7.5%, carotid ultrasound 0%, ECG 32.5%, and pulse wave analysis 12.5%. When broken down by sex, for women, traditional risk factors contribute 37.5%, magnetic resonance imaging 0%, carotid ultrasound 10%, ECG 30%, and pulse wave analysis 22.5% to the top ten. For men, the breakdown is traditional risk factors 40%, magnetic resonance imaging 5%, carotid ultrasound 2.5%, ECG 32.5%, and pulse wave analysis 20%.

## 4 Discussion

### Women are traditionally under-diagnosed for cardiovascular diseases

The lack of sex-specific criteria is one factor contributing to the under-diagnosis of cardiovascular diseases in women compared to men [6]. From Figure 2, we conclude that in the UK Biobank database, women are nearly 2x more under-diagnosed than men for first degree AV block, and 1.4x for dilated cardiomyopathy when using standard sex-neutral criteria. When accounting for average sex differences in PQ interval, left ventricle diameter, and ejection fractions, the fraction by which women are under-diagnosed would increase even further.

For essential primary hypertension, based on the current sex-neutral criteria, women and men are equally under-diagnosed. Yet, as Figure 2 suggests, women have on average lower systolic and diastolic blood pressures than men. If sex-specific criteria were used, women would be under-diagnosed for hypertension. Lastly, women have a smaller wall thickness than men, but the criteria for diagnosing hypertrophic cardiomyopathy are the same. Here, women would again benefit from sex-specific criteria.

### The novel SAINT model outperforms XGBoost in predicting the risk for cardiovascular disease

Using a dataset of UK Biobank patients who underwent cardiovascular clinical tests, we designed sixty classifiers based on relevant features, sex, and disease categories. We compared the new deep learning model SAINT to the state of the art for tabular data, XGBoost, and to an MLP deep learning baseline. We found that SAINT showed the highest cardiovascular disease prediction AUC in nearly every case, XGBoost typically achieved the second-best AUC, and MLP the lowest AUC.

SAINT is specifically designed for tabular data which makes its purpose-driven architecture significantly better for our task than an out-of-the-box MLP. The best performance of the SAINT classifiers can be attributed, at least in part, to the fact that our data are composed of numerical, continuous features, which are known to favor the performance of SAINT over classical approaches, such as XGBoost [23]. The MLP trained with all cardiovascular and Framingham Risk Score features outperformed the state of the art XGBoost method trained with Framingham-only features for all but two dataset variants, which indicates that having access to more features significantly increases model fidelity for this dataset.

To date, the SAINT architecture has not been applied for risk analysis in cardiovascular disease. Its remarkable performance not only holds promise for further clinical studies of both cardiovascular disease and other conditions, but also suggests that deep learning approaches could re-surface as viable methods for tabular clinical data modeling. Since deep learning frameworks require large datasets for effective training, we expect SAINT to improve even more as the size of the available medical data increases.

### Traditional risk factors are not all equally important

Age, body mass index, HDL and total cholesterol, and systolic blood pressure were the most common factors across sex and disease, with smoking status and diabetes status present for men only, as Table 4 suggests. A previous cardiovascular risk prediction study from the UK Biobank used a machine learning pipeline that included 423,604 participants and 473 features, including the Framingham Risk Score, health and medical history, lifestyle and environment, blood assays, physical activity, family history, physical measures, psycho-social factors, dietary and nutritional information, and sociodemographics [19]. This group did not have access to cholesterol levels at the time of their analysis, but they did find that the top feature for men and women was age [19], which we reported as well in Table 4. The previous study reported smoking status and systolic blood pressure in the top ten for both women and men [19], while we found smoking status only for men and central systolic blood pressure for women.

When broken down by disease category, for hypertension measures of blood pressure, body mass index, age, and cholesterol ranked highly. For men, diabetes status was an important feature, but not for women. Interestingly, only total cholesterol, not HDL cholesterol, ranked in the top ten for men, while both appeared for women. For ischemic diseases, age, HDL and total cholesterol, and body mass index were in the top ten for both women and men. For conduction disorders, for women, age and HDL cholesterol were in the top features while for men, age and body mass index appeared. The traditional risk factors in the Framingham Risk Score appear to be most important for a general calculation of cardiovascular risk, but our study suggests to re-evaluate it by taking into account sex- and disease-specific categories.

### ECG recordings are the most effective feature to augment the Framingham Risk Score

For women, traditional risk factors made up 37.5% of the top ten features, while for men, they made up 40%, as we conclude from Table 4. ECG features appeared next in the top 10, making up 30% and 32.5% for women and men, then pulse wave analysis with 22.5% and 20%.

ECG features such the QRS duration, QT duration, and T-wave morphology are associated with increased cardiovascular mortality [63–65]. Women are known to have a shorter PQ interval and QRS duration, longer QTC, and different T-wave morphology than men [64, 66]. All of these features appeared in the top ten from our feature importance analysis across several sex- and disease-categories in Table 4. Adding the ECG features with high SHAP values to the traditional Framingham Risk Score features would be a simple yet effective strategy to increase the predictive potential of cardiovascular disease models.

### Central blood pressure is more predictive of cardiovascular disease risk than brachial blood pressure

Pulse wave analysis provides multiple measures of blood pressure. In multiple sex- and disease-categories in Table 4, central systolic blood pressure ranked higher than systolic brachial blood pressure. Central blood pressure relates closely to the load on the coronary and cerebral arteries and, as such, is more strongly correlated with vascular diseases and negative outcomes than brachial blood pressure [67]. Other pulse wave features like pulse rate, arterial stiffness index, reflection index, and mean arterial pressure that made the top ten in Table 4 have also previously been linked to increased risk of cardiovascular disease [68–71].

### Carotid ultrasounds provide an accessible way to monitor ischemic heart diseases

Carotid ultrasounds measure carotid intima-media thickness; a thicker intima-media thickness may indicate atherosclerosis of the carotid artery leading to the brain [72]. Increasing evidence suggests that atherosclerosis in the carotid artery is associated with atherosclerosis in the coronary artery, leading to increased risk of stroke, myocardial infarction, and other ischemic heart diseases [47, 72, 73]. Because the carotid artery is easily accessible compared to the coronary artery, carotid ultrasounds provide a non-invasive, simple way to screen patients for increased risk of cardiovascular diseases [73]. In Table 4, we found that three features from the carotid ultrasound for women and one for men appeared in the prediction of ischemic diseases. One carotid ultrasound features also appeared for women for the prediction of any cardiovascular disease. As such, carotid ultrasounds provide valuable insight in an individual’s risk for ischemic diseases, regardless of sex, and may be especially useful for monitoring the cardiovascular disease risk of women in general.

### Limitations and future work

Our study provides a first step towards rethinking risk indicators for cardiovascular disease in view of big data and machine learning. While our results provide encouraging evidence of the added value of leveraging both technologies, it is important to be aware of the limitations of our current approach and, ideally, address them in future follow-up studies. First, and most obviously, the outcome of our approach is only as good as the clinical diagnoses that define our classification. It would be highly beneficial, and actually very feasible with modern machine learning techniques, to perform a comprehensive study of human-level error by evaluating expert clinician performance on the utilized datasets. This would provide an estimate of the Bayes error rates for our 12 datasets that could then be compared to the SAINT model performance. Secondly, the population of the UK Biobank is fairly homogeneous, so our classifiers might not be generalizable to participants outside the United Kingdom who are from more diverse racial and ethnic backgrounds. Third, since SAINT was the best-performing approach for our classification tasks, future studies could focus on integrating a comprehensive feature importance pipeline, such as SHAP, into the SAINT model evaluation. This would leverage the high performance of the SAINT method, and could translate to even more informative and credible feature significance rankings. Finally, a direct comparison between the XGBoost and SAINT feature analyses would provide further insight into the sensitivity of feature identification with respect to the specifics of a given learning architecture.

## 5 Conclusion

Women are under-diagnosed for cardiovascular diseases compared to men. Unarguably, there is urgent need for sex-specific diagnostic criteria. Deep learning provides powerful tools to quantify precisely how well traditional risk factors, like the Framingham Risk Score, predict the risk of cardiovascular disease–for females, males, or both sexes combined. Alarmingly, our deep-learning study revealed that, for first-degree atrioventricular block and dilated cardiomyopathy, women are under-diagnosed 2x and 1.4x more than men. Inversely, without much extra work, our deep-learning approach allows us to identify and rank the most predictive features for different types of cardiovascular disease, sex-specifically and sex-neutrally. We found that, out of four commonly used clinical tests–electrocardiograms, magnetic resonance imaging, carotid ultrasound, and pulse wave analysis–electrocardiogram features showed the most promise in increasing cardiovascular disease prediction. A more accurate individualized risk prediction in cardiovascular disease would enable personalized treatment and prevention strategies, a more effective allocation of medical resources, and an early and precise identification of high-risk individuals, towards the ultimate goal to improve patient outcomes, reduce morbidity and mortality, and improve quality of life.

## Data Availability

All data produced in the present work are contained in the manuscript are available on request

## Acknowledgments

This research was conducted using the October 2022 release of the UK Biobank Resource under Application Number 89726. It uses data provided by patients and collected by the National Health Service England as part of their care and support; copyright 2022 National Health Service England; re-used with the permission of the UK Biobank; all rights reserved. This project was done in part for the class CS230 Deep Learning, and we would like to thank Andrew Ng and the teaching assistants for their helpful feedback. This project was supported by an NSF Graduate Research Fellowship to SS, and by the NSF CMMI grants 2318188 and 2320933 and to EK.

## Conflicts of interest

All authors declare no financial competing interests.

## Availability of data and materials

Any researcher can apply to access the UK Biobank to complete health-related research that is in the public interest.

## Code availability

The code for all preprocessing, analyses, and models is available at github.com/LivingMatterLab/CardiovascularDiseaseClassification.

## Authors’ contributions

SS and BK wrote the code, performed the analyses, created the figures, and wrote the manuscript. MP and EK helped design the study and wrote the manuscript. All authors read and approved the final manuscript.

## Appendix A. Computational structure of SAINT architecture

Here we present a brief synthesis of the SAINT framework structure [31]. From a high-dimensional embedding of the input features, a single stage of the SAINT unit computes the output through a series of multi-head self-attention blocks, intersample attention blocks, feed forward layers, and layer normalization layers. Several of these stages are stacked sequentially before the final contextual representation is generated. In each stage, the self-attention block applies attention among the features of a given sample, while the intersample attention applies row-wise attention across different samples for a given feature. Tabular data learning in SAINT is divided into two phases: self-supervised pre-training and supervised fine-tuning. The pre-training phase consists of minimizing the combined contrastive and denoising the cost function without considering the example labels. The fine-tuning phase evaluates a deviation metric between ground truth and model prediction. Throughout this study, we perform a binary classification of cardiovascular disease presence, and use the binary cross-entropy loss function as our deviation metric of choice.

## Appendix B. Hyperparameter tuning

### MPL: A deep learning model for baseline comparison

We apply Bayesian optimization using the Keras tuner to tune the hyperparameters of the multi layer perceptron over the validation data for the dataset with both sexes and any disease. The optimization shows that a multi layer perceptron architecture with 6 hidden layers, 30 units in each hidden layer, and an *L*_2_-regularization parameter *λ* = 10^*−*2^ in each layer provides a robust model performance. We observe that a batch size of 32 yields the fastest convergence, since all datasets are small-to medium-sized. A maximum of 100 epochs allows us to reach convergence, while preserving computational feasibility. We further conclude that the performance of the multi layer perceptron on the validation data is not highly sensitive to changes in other hyperparameters, so we used the default values of the learning rate and the coefficients in the Adam update rule. In principle, we should apply this hyperparameter study to each of the 12 multi layer perceptrons individually. However, the Bayesian tuning process itself is parallelized on a single GPU, and we decided to not tune all 12 multi layer perceptron models individually. We applied the hyperparameters of both sexes, any disease dataset to the remaining 11 models.

### XGBoost: A state of the art analysis of tabular data

Since XGBoost training is significantly faster than multi layer perceptron training and readily parallelizable over the 12 dataset variants, we apply a random search with 5-fold cross-validation to tune the hyperparameters of each of the XGBoost models individually. Specifically, we tune the XGBoost hyperparameters for each of the 12 dataset variants, and for the two input feature groups, all available features (cardiovascular and Framingham Risk Score), and Framingham Risk Score features only. Based on prior work [74, 75], the most valuable hyperparameters to tune include maximum tree depth, learning rate, subsample ratio of the training instances, subsample ratio of columns for each tree, subsampling ratio of columns for each level, and the number of tree estimators. We tune these hyperparameters in a parallelized random search routine. The tuning process minimizes the AUC score over the validation data. We compare two different XGBoost ensembles: one set that has access to all features (cardiovascular and Framingham Risk Score features), and another set that can train on Framingham Risk Score features and body mass index only. Each set of XGBoost models is applied to all 12 dataset variants, and the hyperparameter tuning is performed for each XGBoost model individually. Thus, we compute 24 optimal sets of XGBoost hyperparameters for the resulting 24 population-disease-feature combinations (3 input population groups, 4 disease label sets, 2 feature groups). For evaluation purposes, we consider both the 24 tuned and the 12 untuned XGBoost models separately, which yields a total of 36 XGBoost ensembles for performance analysis. The 12 untuned XGBoost models for the Framingham Risk Score only feature group are not included in the evaluation due to redundancy.

### SAINT: A novel approach for tubular learning

We did not have access to sufficient compute to be able to perform full-scale hyperparameter tuning for the 12 SAINT classifiers. We did, however, make adjustments to hyperparameters that proved to have the most significant effect on the validation performance. Specifically, we increased the weight decay parameter of the AdamW optimization scheme [76] to *w* = 10, since lower values (such as the default of *w* = 10^*−*2^) led to significant overfitting on the training set. We also found that the default choices of the learning rate and other AdamW optimization parameters provided the best validation set performance.

## Notes

### Competing Interest Statement

The authors have declared no competing interest.

### Clinical Protocols

https://github.com/LivingMatterLab/CardiovascularDiseaseClassification

### Author Declarations

This research was conducted using the October 2022 release of the UK Biobank Resource under Application Number 89726. It uses data provided by patients and collected by the National Health Service England as part of their care and support; copyright 2022 National Health Service England; re-used with the permission of the UK Biobank; all rights reserved.

### Summary of Updates

Update of the acknowledgements by request of the UK Biobank from previously "This research was conducted using the UK Biobank Resource under Application Number 89726. The data used were pulled from the October 2022 release of the UK Biobank database." to now: "This research was conducted using the October 2022 release of the UK Biobank Resource under Application Number 89726. It uses data provided by patients and collected by the National Health Service England as part of their care and support; copyright 2022 National Health Service England; re-used with the permission of the UK Biobank; all rights reserved."

